# Evaluating the number of unvaccinated people needed to exclude to prevent SARS-CoV-2 transmissions

**DOI:** 10.1101/2021.12.08.21267162

**Authors:** Aaron Prosser, Bartosz Helfer, David L. Streiner

**Affiliations:** Department of Psychiatry and Behavioural Neurosciences, McMaster University, Hamilton, Ontario, Canada; Meta-Research Centre, University of Wroclaw, Wroclaw, Poland; National Heart and Lung Institute, Imperial College London, London, UK; Department of Psychiatry and Behavioural Neurosciences McMaster University, Hamilton, Ontario, Canada

## Abstract

**Background:** Vaccine mandates and vaccine passports (VMVP) for SARS-CoV-2 are thought to be a path out of the pandemic by increasing vaccination through coercion and excluding unvaccinated people from different settings because they are viewed as being at significant risk of transmitting SARS-CoV-2. While variants and waning efficacy are relevant, SARS-CoV-2 vaccines reduce the risk of infection, transmission, and severe illness/hospitalization in adults. Thus, higher vaccination levels are beneficial by reducing healthcare system pressures and societal fear. However, the benefits of excluding unvaccinated people are unknown.

**Methods:** A method to evaluate the benefits of excluding unvaccinated people to reduce transmissions is described, called the *number needed to exclude* (NNE). The NNE is analogous to the *number needed to treat* (NNT=1/ARR), except the absolute risk reduction (ARR) is the baseline transmission risk in the population for a setting (e.g., healthcare). The rationale for the NNE is that exclusion removes *all* unvaccinated people from a setting, such that the ARR is the baseline transmission risk for that type of setting, which depends on the secondary attack rate (SAR) typically observed in that type of setting and the baseline infection risk in the population. The NNE is the number of unvaccinated people who need to be excluded from a setting to prevent one transmission event from unvaccinated people in that type of setting. The NNE accounts for the transmissibility of the currently dominant Delta (B.1.617.2) variant to estimate the minimum NNE in six types of settings: households, social gatherings, casual close contacts, work/study places, healthcare, and travel/transportation. The NNE can account for future potentially dominant variants (e.g., Omicron, B.1.1.529). To assist societies and policymakers in their decision-making about VMVP, the NNEs were calculated using the current (mid-to-end November 2021) baseline infection risk in many countries.

**Findings:** The NNEs suggest that at least 1,000 unvaccinated people likely need to be excluded to prevent one SARS-CoV-2 transmission event in most types of settings for many jurisdictions, notably Australia, California, Canada, China, France, Israel, and others. The NNEs of almost every jurisdiction examined are well within the range of the NNTs of acetylsalicylic acid (ASA) in primary prevention of cardiovascular disease (CVD) (≥ 250 to 333). This is important since ASA is not recommended for primary prevention of CVD because the harms outweigh the benefits. Similarly, the harms of exclusion may outweigh the benefits. These findings depend on the accuracy of the model assumptions and the baseline infection risk estimates.

**Conclusions:** Vaccines are beneficial, but the high NNEs suggest that excluding unvaccinated people has negligible benefits for reducing transmissions in many jurisdictions across the globe. This is because unvaccinated people are likely *not* at significant risk – in absolute terms – of transmitting SARS-CoV-2 to others in most types of settings since current baseline transmission risks are negligible. Consideration of the harms of exclusion is urgently needed, including staffing shortages from losing unvaccinated healthcare workers, unemployment/unemployability, financial hardship for unvaccinated people, and the creation of a class of citizens who are not allowed to fully participate in many areas of society.

**Registration:** CRD42021292263

**Funding:** This study received no grant from any funding agency, commercial, or not-for-profit sectors. It has also received no support of any kind from any individual or organization. BH is supported by a personal research grant from the University of Wroclaw within the “Excellence Initiative – Research University” framework and by a scholarship from the Polish Ministry of Education and Science. None of these institutions were involved in this research and did not fund it directly.

**Competing interests:** The authors have no competing interests to declare.

**Ethical approval:** Not applicable. All the work herein was performed using publicly available data.

**Data reporting:** The data used in this work are available at https://tinyurl.com/4m8mm4jh and https://decision-support-tools.com/.

## INTRODUCTION

A key global priority for the pandemic caused by severe acute respiratory syndrome coronavirus 2 (SARS-CoV-2) is to return to normality as quickly as possible. This is because many societies are losing their political, economic, psychological, and social tolerance for lockdowns and restrictions, especially since the development of SARS-CoV-2 vaccines. There is growing pressure to implement vaccine mandates and vaccine passports (VMVP) domestically and for international travel as a “ticket” for full participation in post-pandemic life. Consequently, more governments, healthcare organizations, educational institutions, and businesses are adopting VMVP. VMVP are thought to be a path out of the pandemic by (i) increasing vaccination levels through coercion and (ii) excluding those who remain unvaccinated from many types of settings (e.g., work, social gatherings). A central legal justification for exclusion is that unvaccinated people are viewed as being at significant risk of transmitting SARS-CoV-2, which is potentially the most serious externalized risk of the choice to remain unvaccinated.^1^

The benefits of vaccination in response to VMVP are clear. Meta-analyses show that SARS-CoV-2 vaccines are effective at reducing the risk of infection, transmission, and severe illness/hospitalization in adults.^2 3^ Therefore, while variants and waning efficacy may moderate these benefits, higher vaccination levels are beneficial because they will reduce the burden on healthcare systems and reduce societal fear. On the other hand, the benefits of excluding unvaccinated people to reduce SARS-CoV-2 transmissions has not been evaluated quantitatively. This evaluation is a critical first step to determining the costs vs. benefits of VMVP. It is also urgently needed because an unknown percentage of people will not get vaccinated despite VMVP. This means those unvaccinated people will be excluded from many areas of modern life, such as many places of work, leisure, transportation, and education. As of December 2021, this is already happening in many jurisdictions globally.

Therefore, we performed a pre-planned analysis of a larger study of VMVP (CRD42021292263). The purpose of this analysis was to describe a method to evaluate the benefits of excluding unvaccinated people to reduce transmissions analogous to the *number needed to treat* (NNT),^4^ which we call the *number needed to exclude* (NNE). The NNT is 1 divided by the absolute risk reduction (ARR). The NNT is the number of people who need to receive a treatment to prevent one outcome (e.g., myocardial infarction). The difference is that the ARR of the NNE is the baseline transmission risk in the population for a given type of setting (NNE=1/ARR). The rationale for the NNE is that exclusion removes *all* unvaccinated people from a setting (e.g., healthcare), such that the ARR is the baseline transmission risk in that type of setting. The ‘treatment’ is the public health intervention of exclusion via VMVP.

The baseline transmission risk is the probability in the general population that an unvaccinated person is currently infected and transmits SARS-CoV-2 in a given type of setting. In other words, it is the current probability in the general population of a transmission event in a setting from an unvaccinated person. The reciprocal of this probability is the NNE. The baseline transmission risk is estimated by taking the combined probability of the secondary attack rate (SAR) typically observed in that type of setting and the baseline infection risk in the general population. The combined probability is needed to estimate this probability because an unvaccinated person must be infected first before they can transmit SARS-CoV-2. Therefore, the NNE is the number of unvaccinated people who need to be excluded from a setting to prevent one transmission event from unvaccinated people in that type of setting. The NNE pertains to one transmission event, which may include one or more secondary infections, because the SAR is the infection risk among the contacts of the unvaccinated index case. The NNE concerns the direct transmission caused by the unvaccinated person, not subsequent generations of transmission.

The baseline infection risk is the current point-prevalence of infectious cases in the general population. The numerator is the total number of new and existing infectious cases, and the denominator is the population size. It is the ‘baseline’ risk because it is the current risk of getting infected with SARS-CoV-2 in a population. It is the estimated risk that an unvaccinated person gets infected. Point-prevalence is a more appropriate metric of baseline infection risk than incidence or period prevalence for three reasons. First, the population-level risk of getting infected depends not just on new infectious cases, but existing ones too. Second, while risk over time is important, people are primarily concerned about the risk of getting infected with SARS-CoV-2 *right now*, not, for example, over the last 3 months. Second, incidence depends on time at risk. Shorter periods will have a lower incidence than longer periods. There is no non-arbitrary way of defining the “correct” time at risk. Point-prevalence does not have this arbitrariness because it uses the current prevalence. Like the NNT, *time* is implicit in the NNE since it relates to the period over which the risk is measured. The NNE relates to the current risk because it is based on the current point-prevalence of infectious cases and thus the current baseline transmission risk. In practice, the risk would be over one day given how point-prevalence data is typically reported. Time is also incorporated into the NNE when one calculates the NNEs over time using changing baseline infection risks.

Critically, societies need to be mindful of the baseline infection risk because when it is low, baseline transmission risks are also low, which exponentially increases the NNE (Figure 1). Figure 1 shows that the transmission risk reduction gained from excluding unvaccinated people becomes negligible when baseline transmission risks are low, especially below 0.40% to 0.30%. This is the range of the ARRs of acetylsalicylic acid (ASA) in primary prevention of cardiovascular disease (CVD) (NNTs ≥ 250 to 333).^5-7^ Notably, ASA is not recommended for primary prevention in all adults because the negligible ARRs do not outweigh the risk of harm.^7^ Similarly, the harms of exclusion via VMVP may outweigh the benefits when baseline infection risks are low.

**Figure 1.**
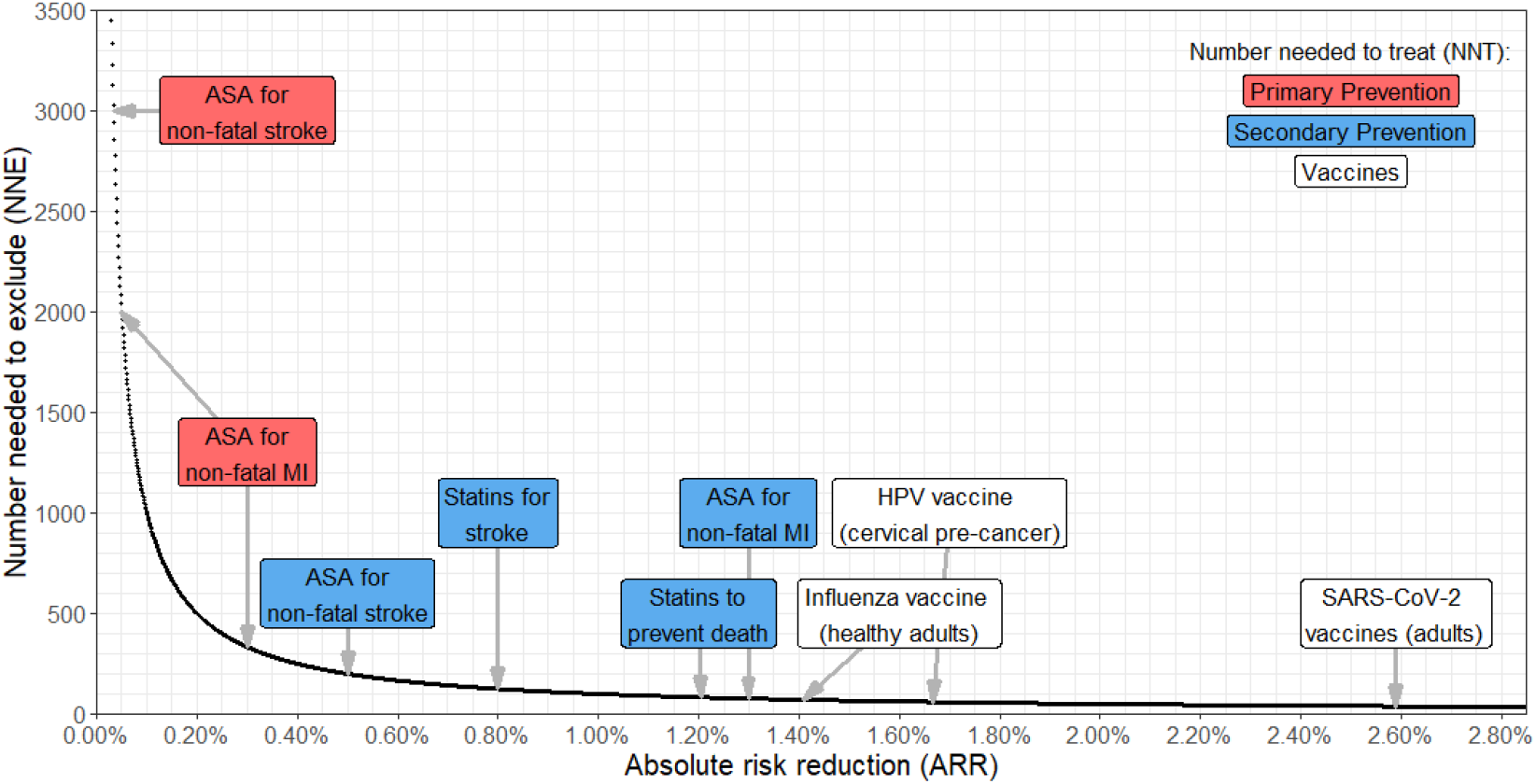
Relationship between baseline transmission risk and the number needed to exclude (NNE). The number needed to treat (NNT) has the same scale as the NNE (NNT=1/ARR). The NNTs of other interventions are shown for comparison.^3 5 6 8-11^ The ARRs of the influenza and SARS-CoV-2 vaccines are for reducing the risk of infection. The ARR of the HPV vaccine is for preventing any cervical pre-cancer. ARR=absolute risk reduction. MI=myocardial infarction. HPV=human papillomavirus.

The NNE was calculated for six types of settings: (i) *households*, (ii) *social gatherings* (e.g., meals, conversations, social gatherings with friends/family), (iii) *casual close contacts* (e.g., public areas or buildings), (iv) *work or study places*, (v) *healthcare*, and (vi) *travel/transportation*. The NNEs accounted for the transmissibility of the Delta (B.1.617.2) variant because it currently predominates in most countries. The NNE can account for the increased transmissibility of future potentially dominant variants (e.g., Omicron, B.1.1.529). First, the NNEs for each type of setting were modeled as a function of simulated baseline infection risks (0.10% to 100%). Second, to assist societies and policymakers in their decision-making about excluding unvaccinated people via VMVP, the NNEs were calculated for each type of setting using the current (mid-to-end November 2021) baseline infection risk in many countries around the globe.

## METHODS

The NNE for the *i*th type of setting was modeled as follows:

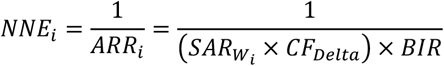

The ARR of excluding unvaccinated people from the *i*th setting is the baseline transmission risk for that type of setting. The baseline transmission risk is estimated by multiplying robust estimates of the wild-type SAR for the *i*th setting 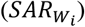 by a correction factor for the increased transmissibility of the Delta variant over the wild-type (*CF*_*Delta*_) and the baseline infection risk (*BIR*) in the population. *CF*_*Delta*_ is necessary because our robust SARs were derived from meta-analyses of studies from 2020 when the wild-type form of SARS-CoV-2 predominated, whereas the Delta variant currently predominates in most counties. We needed to use the wild-type SARs because studies of the SARs of variants occurred during the vaccine period (2021), which would add a confounding factor to the model. *CF*_*Delta*_ was 1.97 in the model because a global analysis of effective reproduction numbers found that the Delta variant was 97% more transmissible than the wild-type.^12^ The model can account for future potentially dominant variants (e.g., Omicron, B.1.1.529) as data emerges regarding its transmissibility over the wild-type by changing the value of *CF*. The corrected robust SARs 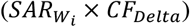 must be multiplied by the *BIR* because SARs are the transmission risks *among infected people*, not the general population. Estimating the transmission risk in the general population requires calculating the combined probability of infection and transmission because a person must be infected first before they can transmit SARS-CoV-2. To model how the NNE for the *i*th setting changes as a function of the baseline infection risk, we simulated values of *BIR* ranging from 0.10% to 100% and plotted the results.

Robust estimates of the wild-type SARs 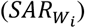 were obtained by calculating the mean and 95% confidence intervals (CIs) of meta-analytic estimates of the SARs for the six types of settings mentioned above. Social gatherings are high intensity contacts (e.g., gatherings of friends/family), whereas casual close contacts are lower intensity contacts (e.g., contact in a public area or building). The 95% CIs of the NNEs were calculated from the upper and lower limits of these robust SARs. To identify these meta-analyses, we searched PubMed from inception to November 26, 2021, with no language restrictions. We used the search terms “SARS-CoV-2” or “COVID-19” combined with “secondary attack*” and “meta-analysis”. Two authors (AP, BH) screened the titles/abstracts to determine potential eligibility. Full-texts of potentially eligible reports were retrieved and assessed to determine eligibility. Data were independently extracted. Disagreements at either the screening, full-text review, or data extraction stages were resolved through consensus between the two authors. Reports were included if they were a meta-analysis of pre-vaccine studies (2020) of SARs and estimated the SAR for at least one of the six types of settings. Meta-analyses of studies during the vaccine period (2021) were excluded to remove the confounding influence of vaccination on SARs. Random effect model data were extracted whenever possible. 13 citations were identified from the search. 1 meta-analysis^13^ was identified in our pilot study.^3^ The meta-analytic estimates of the SARs were extracted from the 7 meta-analyses meeting the inclusion criteria.^13-19^ All analyses were conducted in R.^20^

The NNE model makes four assumptions. First, we assume the Delta variant correction factor (*CF*_*Delta*_=1.97) is accurate. We checked this assumption by comparing the model predictions with the observed SARs of the Delta variant in other studies (see Results). Second, the model assumes the SARs for each setting type are relatively consistent because of systematic differences in the intensity of contact (proximity, duration of contact, use of precautions, e.g., masking). Lower contact intensity settings should have lower SARs than higher contact intensity settings. For instance, we assume that households will consistently have higher SARs than healthcare settings because the intensity of contact is higher, which is suggested by the fact that households are a major driver of SARS-CoV-2 transmissions.^16^ We supported this assumption by utilizing meta-analytic estimates in order to arrive at robust estimates of the central tendency and distribution of SARs for each type of setting.

Third, the model assumes the baseline infection risk is relatively stable across individuals in a population. Given that there will inevitably be local variation in risk, we addressed this assumption by modeling a range of baseline infection risks from 0.10% to 100% and plotted the 95% CIs around the NNEs based on the 95% CIs of the corrected robust SARs. Similarly, we calculated the 95% CIs around the NNEs for each jurisdiction we examined (see below) using the 95% CI of the baseline infection risk in the jurisdiction.

Fourth, the model assumes (i) unvaccinated people have no natural immunity and (ii) the contacts of infected unvaccinated index cases have no vaccine or natural immunity. This is because the SARs were derived from the pre-vaccine period. It was not possible to know from the meta-analyses to what extent the studies included participants with a history of prior infection with SARS-CoV-2. However, it is reasonable to assume that the vast majority had no natural immunity in these studies. This is because a global meta-analysis of studies from 2020 estimated the median seroprevalence of SARS-CoV-2 antibodies in the general population across countries was 4.5% (IQR: 2.4%, 8.4%).^21^ This is important because populations with higher levels of vaccine and/or natural immunity will have lower SARs and thus lower baseline transmission risks, regardless of the baseline infection risk in the population. Accordingly, higher levels of vaccine/natural immunity in a population will increase the NNE, independent of the current baseline infection risk. By assuming this, the corrected robust SARs 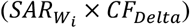 are the worst-case scenario because they are higher than what would be predicted in a population with some level of vaccine/natural immunity. This is a strong conservative assumption since many jurisdictions already have high levels of vaccine/natural immunity (as of December 2021). Vaccine/natural immunity in a population will reduce the denominator of the NNE because they will reduce the value of 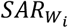, which will reduce the baseline transmission risk, and thus increase the NNE. Therefore, the calculated NNEs likely represent the *minimum* number of unvaccinated people needed to exclude to prevent one SARS-CoV-2 transmission event in each type of setting.

Research supports this impact of vaccine/natural immunity on the baseline transmission risk in a population. A large Belgian study of the period of Alpha (B.1.1.7) predominance found that natural immunity had the same effectiveness (∼50-60%) for reducing the SAR among unvaccinated close contacts as full vaccination with BNT162b2 and mRNA-1273.^22^ It was also the same (∼75-85%) for contacts with natural immunity vs. vaccine immunity who were exposed to unvaccinated index cases. A large Dutch study during the same period found that the relative risk reduction (RRR) is 65% when unvaccinated index cases had contact with fully vaccinated (SAR=11%) vs. unvaccinated (SAR=31%) household members.^23^ In summary, this research shows that if an unvaccinated index case has natural immunity or their contacts have either vaccine or natural immunity, the SAR is reduced for the contacts of infected unvaccinated people. Therefore, vaccine/natural immunity in a population will reduce the baseline transmission risks in that population for any given setting, and thus increase the NNE.

One can incorporate the effect of vaccine/natural immunity into the NNE model by adding additional correction factors to the corrected robust SARs 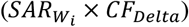. These correction factors can be estimated using the RRRs of vaccine/natural immunity on SARs in unvaccinated index cases and their contacts, each weighted by the proportion of the population fully vaccinated and with natural immunity. However, we opted not to do this for two reasons. First, we wanted to make as few assumptions as possible so that the model remained close to the data. Second, we wanted to produce conservative estimates of the NNEs for each setting.

The NNEs were also calculated using estimates of current baseline infection risks and their 95% CIs for a sample of countries across the globe. The same computations above were used, except the 95% CIs of the NNEs were calculated from the upper and lower limits of the baseline infection risks. Estimates of the current point-prevalence of infectious cases in these jurisdictions were taken from a global database (https://decision-support-tools.com/) developed and regularly updated by Defence Research and Development Canada (DRDC), an agency of the Canadian Department of National Defence. Its estimation methods are detailed online. Australia had data on its six states, the Australian Capital Territory, and Northern Territory. China had data on its provinces, municipalities, and autonomous regions except Tibet. For both countries, these data were extracted and means were used to estimate national-level risks. No data for Sweden was available for the time point we extracted. Regional data for Canada and the United States of America (USA) were available, extracted, and means were used to estimate state/provincial-level and national-level risks. We extracted the most recent dataset posted online on November 26^th^, 2021. 71% of the dataset was from November 21^st^ to 25^th^, 2021, 29% were from November 7^th^ to 11^th^, 2021. Thus, point-prevalence data were from mid-to-end November 2021.

## RESULTS

Table 1 displays the wild-type vs. corrected robust SARs for each type of setting. The predicted household SAR for the Delta variant (32.59%, 95% CI: 27.04%, 38.14%) was 26% higher than the observed SARs pooled across recent studies of unvaccinated index cases infected with the Delta variant who had contact with primarily household members (25.90%, 95% CI: 19.02%, 32.78%).^24-26^ In other words, the Delta variant correction factor (1.97) based on the effective reproduction numbers was an accurate but conservative assumption because it increased the SARs more than what has been observed in the real-world. This additional conservative assumption of the model is another reason why the NNEs are likely the minimum NNEs.

**Table 1.** Robust estimates of the secondary attack rates to model the NNE for each setting

| Type of setting | Robust SARs | Corrected robust SARs |
| --- | --- | --- |
| | (Wild-type: $SAR_{Wt}$) (95% CI) | (Delta variant: $SAR_{Wt} \times CF_{Delta}$) (95% CI) |
| Households | 16.54% (13.73%, 19.36%) | 32.59% (27.04%, 38.14%) |
| Social gatherings | 5.93% (5.87%, 6.00%) | 11.69% (11.56%, 11.82%) |
| Casual close contacts | 1.55% (0.86%, 2.24%) | 3.05% (1.70%, 4.40%) |
| Work/study places | 1.47% (0.96%, 1.98%) | 2.89% (1.88%, 3.89%) |
| Healthcare | 1.50% (0.12%, 2.88%) | 2.96% (0.24%, 5.67%) |
| Travel/transportation | 2.23% (0.00%, 4.95%) | 4.40% (0.00%, 9.75%) |

95% CI=95% confidence interval. Brackets are the 95% confidence intervals (lower limit, upper limit). SAR=secondary attack rate. NNE=number needed to exclude. Note, the CIs were widest for travel/transportation and because the lower limit crossed zero it was set to zero. $CF_{Delta}=1.97$ . See Methods for details.

Figure 2 plots the NNEs for the six types of settings using simulated baseline infection risks. These NNEs can be interpreted in terms of the NNTs of ASA in primary prevention of CVD (Figure 1). NNEs within the range of the NNTs of ASA in primary prevention of CVD (≥ 250 to 333) are considered ‘high’ because it means the benefits of excluding unvaccinated people to reduce transmission risks are negligible, much like the ARRs of ASA are negligible in preventing myocardial infarction and strokes in people with no history of CVD.^5-7^ These negligible ARRs result in high NNTs and are the reason why ASA is not recommended for primary prevention of CVD in all adults since the benefits do not outweigh the risk of harm.^7^ As seen in Figure 2, for household settings, NNEs start to become high when baseline infection risks are *≤* 1%. For social gatherings, NNEs start to become high when baseline infection risks are *≤* 3%. For casual close contacts, work/study places, and healthcare settings, NNEs start to become high when baseline infection risks are *≤* 10%. For travel/transportation settings, NNEs start to become high when baseline infection risks are *≤* 5-10%. Figure 2 shows that when baseline infection risks are below 1%, baseline transmission risks become very small, such that thousands of unvaccinated people likely need to be excluded from most types of settings to prevent one SARS-CoV-2 transmission event. High NNEs mean that, when the current point-prevalence of infectious cases in the population is below these thresholds, unvaccinated people are likely *not* at significant risk – in absolute terms – of transmitting SARS-CoV-2 to others.

**Figure 2.**
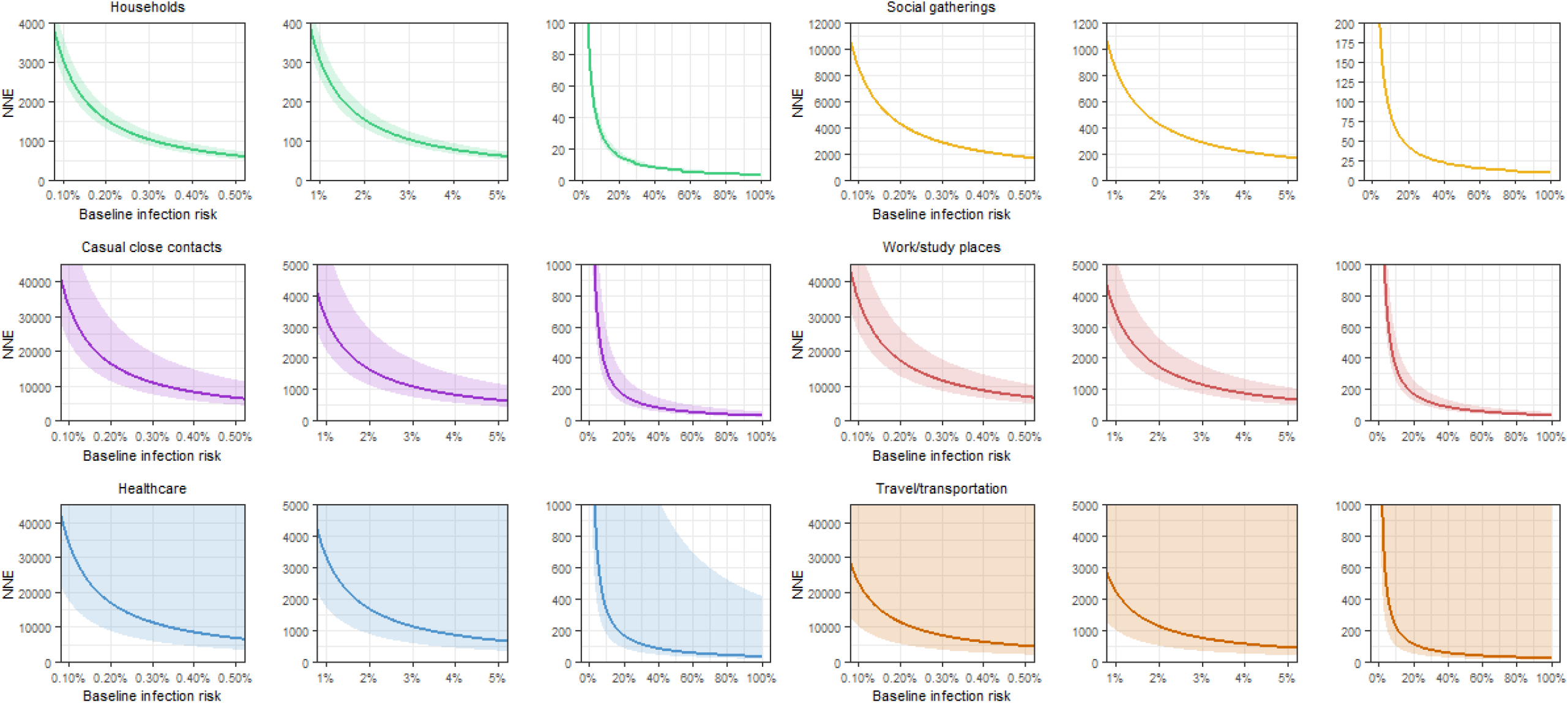
Modeling the number of unvaccinated people needed to exclude (NNE) from different types of settings to prevent one SARS-CoV-2 transmission event in that setting. The NNEs are plotted as a function of simulated baseline infection risks (0.10% to 100%) and corrected for the increased transmissibility of the Delta variant. To better visualize the NNEs at different baseline infection risks, each setting is plotted from 0.10% to 0.50%, 1% to 5%, and 0% to 100%.

Table 2 displays the NNEs for many jurisdictions from the DRDC database estimates of the baseline infection risk as of mid-to-end November 2021. Many of these jurisdictions have implemented some form of VMVP resulting in the exclusion of unvaccinated people from different areas of these societies. NNEs are color-coded by the lower limit of the 95% CI: yellow (≥ 1,000 and *<* 10,000), orange (≥ 10,000 and *<* 100,000), red (≥ 100,000 and *<* 1,00,000), and black (≥ 1,000,000). NNEs ≥ 1,000,000 were truncated to ≥ 1M. The NNEs of almost every jurisdiction and setting type are well within the range of the NNTs of ASA in primary prevention of CVD (≥ 250 to 333). The NNEs suggest that at least 1,000 unvaccinated people likely need to be excluded to prevent one SARS-CoV-2 transmission event in most types of settings for many jurisdictions, notably Australia, California, Canada, China, France, Israel, and others. Given the conservative assumptions of the model, these are likely the minimum NNEs.

**Table 2.** Global estimates of the number of unvaccinated people needed to exclude (NNE) to prevent one SARS-CoV-2 transmission event

|  |  |  |  |  |  |  |  |
| --- | --- | --- | --- | --- | --- | --- | --- |
| Jurisdiction | Estimated baseline<br>infection risks as of mid-to-end November 2021†<br>(95% CI) | Estimated number of unvaccinated people needed to exclude (NNE)<br>to prevent one SARS-CoV-2 transmission event in each type of setting (95% CI) |  |  |  |  |  |
|  |  | Households | Social gatherings | Casual close contacts | Work/study places | Healthcare | Travel/<br>transportation |
| Argentina | 0.0798%<br>(0.0663%, 0.1091%) | 3,843<br>(2,813, 4,626) | 10,716<br>(7,842, 12,898) | 41,019<br>(30,019, 49,373) | 43,349<br>(31,724, 52,178) | 42,386<br>(31,019, 51,019) | 28,468<br>(20,834, 34,266) |
| Australia | 0.2391%<br>(0.135%, 0.3425%) | 1,283<br>(896, 2,273) | 3,578<br>(2,498, 6,338) | 13,695<br>(9,562, 24,262) | 14,473<br>(10,106, 25,640) | 14,151<br>(9,881, 25,071) | 9,504<br>(6,637, 16,839) |
| Australian Capital Territory | 0.2908%<br>(0.1495%, 0.5032%) | 1,055<br>(610, 2,052) | 2,942<br>(1,700, 5,722) | 11,262<br>(6,508, 21,904) | 11,901<br>(6,878, 23,149) | 11,637<br>(6,725, 22,634) | 7,816<br>(4,517, 15,202) |
| New South Wales | 0.0903%<br>(0.0614%, 0.1252%) | 3,397<br>(2,450, 4,998) | 9,471<br>(6,832, 13,934) | 36,255<br>(26,153, 53,341) | 38,315<br>(27,639, 56,371) | 37,464<br>(27,025, 55,119) | 25,162<br>(18,151, 37,020) |
| Northern Territory | 0.3234%<br>(0.1742%, 0.6011%) | 949<br>(510, 1,761) | 2,645<br>(1,423, 4,910) | 10,125<br>(5,448, 18,795) | 10,700<br>(5,757, 19,863) | 10,463<br>(5,629, 19,421) | 7,027<br>(3,781, 13,044) |
| Queensland | 0.0003%<br>(0.0001%, 0.001%) | ≥ 1M<br>(320,491, ≥ 1M) | ≥ 1M<br>(893,569, ≥ 1M) | ≥ 1M<br>(≥ 1M, ≥ 1M) | ≥ 1M<br>(≥ 1M, ≥ 1M) | ≥ 1M<br>(≥ 1M, ≥ 1M) | ≥ 1M<br>(≥ 1M, ≥ 1M) |
| South Australia | 0.0002%<br>(0.0001%, 0.0005%) | ≥ 1M<br>(606,215, ≥ 1M) | ≥ 1M<br>(≥ 1M, ≥ 1M) | ≥ 1M<br>(≥ 1M, ≥ 1M) | ≥ 1M<br>(≥ 1M, ≥ 1M) | ≥ 1M<br>(≥ 1M, ≥ 1M) | ≥ 1M<br>(≥ 1M, ≥ 1M) |
| Tasmania | 0.0009%<br>(0.0003%, 0.002%) | 337,842<br>(157,044, 902,176) | 941,943<br>(437,857, ≥ 1M) | ≥ 1M<br>(≥ 1M, ≥ 1M) | ≥ 1M<br>(≥ 1M, ≥ 1M) | ≥ 1M<br>(≥ 1M, ≥ 1M) | ≥ 1M<br>(≥ 1M, ≥ 1M) |
| Victoria | 1.2068%<br>(0.694%, 1.5058%) | 254<br>(204, 442) | 709<br>(568, 1,233) | 2,714<br>(2,175, 4,719) | 2,868<br>(2,298, 4,987) | 2,804<br>(2,247, 4,876) | 1,883<br>(1,509, 3,275) |
| Western Australia | 0.0004%<br>(0.0002%, 0.001%) | 817,673<br>(292,777, ≥ 1M) | ≥ 1M<br>(816,298, ≥ 1M) | ≥ 1M<br>(≥ 1M, ≥ 1M) | ≥ 1M<br>(≥ 1M, ≥ 1M) | ≥ 1M<br>(≥ 1M, ≥ 1M) | ≥ 1M<br>(≥ 1M, ≥ 1M) |
| Brazil | 0.458%<br>(0.302%, 0.7268%) | 670<br>(422, 1,016) | 1,868<br>(1,177, 2,833) | 7,151<br>(4,506, 10,846) | 7,557<br>(4,762, 11,462) | 7,389<br>(4,656, 11,207) | 4,963<br>(3,127, 7,527) |
| Canada | 1.1221%<br>(0.7015%, 1.7193%) | 273<br>(178, 437) | 762<br>(498, 1,220) | 2,919<br>(1,905, 4,669) | 3,085<br>(2,013, 4,934) | 3,016<br>(1,968, 4,824) | 2,026<br>(1,322, 3,240) |
| Alberta | 0.6645%<br>(0.356%, 1.2135%) | 462<br>(253, 862) | 1,288<br>(705, 2,403) | 4,929<br>(2,699, 9,200) | 5,209<br>(2,852, 9,722) | 5,093<br>(2,789, 9,506) | 3,421<br>(1,873, 6,385) |
| British Columbia | 1.0978%<br>(0.7027%, 1.5806%) | 280<br>(194, 437) | 779<br>(541, 1,217) | 2,983<br>(2,072, 4,660) | 3,153<br>(2,190, 4,925) | 3,083<br>(2,141, 4,816) | 2,070<br>(1,438, 3,235) |
| Manitoba | 2.2159%<br>(1.3999%, 3.2252%) | 138<br>(95, 219) | 386<br>(265, 611) | 1,478<br>(1,015, 2,339) | 1,562<br>(1,073, 2,472) | 1,527<br>(1,049, 2,417) | 1,026<br>(705, 1,624) |
| New Brunswick | 1.2565%<br>(0.8438%, 1.7817%) | 244<br>(172, 364) | 681<br>(480, 1,014) | 2,606<br>(1,838, 3,881) | 2,755<br>(1,943, 4,102) | 2,693<br>(1,899, 4,011) | 1,809<br>(1,276, 2,694) |
| Newfoundland and Labrador | 0.034%<br>(0.0147%, 0.0639%) | 9,030<br>(4,804, 20,878) | 25,178<br>(13,395, 58,212) | 96,381<br>(51,275, 222,832) | 101,857<br>(54,188, 235,492) | 99,593<br>(52,984, 230,259) | 66,891<br>(35,586, 154,652) |
| Northwest Territories | 2.0711%<br>(1.0044%, 3.4395%) | 148<br>(89, 305) | 413<br>(249, 852) | 1,581<br>(952, 3,260) | 1,671<br>(1,006, 3,446) | 1,634<br>(984, 3,369) | 1,097<br>(661, 2,263) |
| Nova Scotia | 0.3008%<br>(0.1791%, 0.4943%) | 1,020<br>(621, 1,714) | 2,844<br>(1,731, 4,778) | 10,888<br>(6,625, 18,290) | 11,507<br>(7,001, 19,329) | 11,251<br>(6,846, 18,899) | 7,557<br>(4,598, 12,694) |
| *Nunavut* | 0.0000%<br>(0.0000%, 0.0011%) | ≥ 1M<br>(269,662, ≥ 1M) | ≥ 1M<br>(751,850, ≥ 1M) | ≥ 1M<br>(≥ 1M, ≥ 1M) | ≥ 1M<br>(≥ 1M, ≥ 1M) | ≥ 1M<br>(≥ 1M, ≥ 1M) | ≥ 1M<br>(≥ 1M, ≥ 1M) |
| *Ontario* | 0.9107%<br>(0.5362%, 1.4796%) | 337<br>(207, 572) | 939<br>(578, 1,595) | 3,596<br>(2,213, 6,107) | 3,801<br>(2,339, 6,454) | 3,716<br>(2,287, 6,311) | 2,496<br>(1,536, 4,239) |
| *Prince Edward Island* | 0.7575%<br>(0.4458%, 1.0521%) | 405<br>(292, 688) | 1,129<br>(813, 1,919) | 4,323<br>(3,113, 7,346) | 4,569<br>(3,290, 7,764) | 4,468<br>(3,217, 7,591) | 3,001<br>(2,160, 5,099) |
| *Quebec* | 1.611%<br>(1.0858%, 2.3744%) | 190<br>(129, 283) | 531<br>(360, 788) | 2,033<br>(1,379, 3,016) | 2,148<br>(1,458, 3,187) | 2,101<br>(1,425, 3,117) | 1,411<br>(957, 2,093) |
| *Saskatchewan* | 0.6956%<br>(0.3614%, 1.1382%) | 441<br>(270, 849) | 1,230<br>(752, 2,367) | 4,708<br>(2,877, 9,061) | 4,976<br>(3,041, 9,576) | 4,865<br>(2,973, 9,363) | 3,268<br>(1,997, 6,289) |
| *Yukon* | 9.2998%<br>(6.5432%, 13.4933%) | 33<br>(23, 47) | 92<br>(63, 131) | 352<br>(243, 501) | 372<br>(256, 529) | 364<br>(251, 517) | 244<br>(168, 347) |
| **China** | 0.0002%<br>(0.0001%, 0.0005%) | ≥ 1M<br>(681,427, ≥ 1M) | ≥ 1M<br>(≥ 1M, ≥ 1M) | ≥ 1M<br>(≥ 1M, ≥ 1M) | ≥ 1M<br>(≥ 1M, ≥ 1M) | ≥ 1M<br>(≥ 1M, ≥ 1M) | ≥ 1M<br>(≥ 1M, ≥ 1M) |
| *Anhui* | 0.0000%<br>(0.0000%, 0.0000%) | Inf<br>(Inf, Inf) | Inf<br>(Inf, Inf) | Inf<br>(Inf, Inf) | Inf<br>(Inf, Inf) | Inf<br>(Inf, Inf) | Inf<br>(Inf, Inf) |
| *Beijing* | 0.0001%<br>(0.0000%, 0.0002%) | ≥ 1M<br>(≥ 1M, ≥ 1M) | ≥ 1M<br>(≥ 1M, ≥ 1M) | ≥ 1M<br>(≥ 1M, ≥ 1M) | ≥ 1M<br>(≥ 1M, ≥ 1M) | ≥ 1M<br>(≥ 1M, ≥ 1M) | ≥ 1M<br>(≥ 1M, ≥ 1M) |
| *Chongqing* | 0.0000%<br>(0.0000%, 0.0000%) | ≥ 1M<br>(≥ 1M, ≥ 1M) | ≥ 1M<br>(≥ 1M, ≥ 1M) | ≥ 1M<br>(≥ 1M, ≥ 1M) | ≥ 1M<br>(≥ 1M, ≥ 1M) | ≥ 1M<br>(≥ 1M, ≥ 1M) | ≥ 1M<br>(≥ 1M, ≥ 1M) |
| *Fujian* | 0.0000%<br>(0.0000%, 0.0000%) | ≥ 1M<br>(≥ 1M, ≥ 1M) | ≥ 1M<br>(≥ 1M, ≥ 1M) | ≥ 1M<br>(≥ 1M, ≥ 1M) | ≥ 1M<br>(≥ 1M, ≥ 1M) | ≥ 1M<br>(≥ 1M, ≥ 1M) | ≥ 1M<br>(≥ 1M, ≥ 1M) |
| *Gansu* | 0.0000%<br>(0.0000%, 0.0000%) | ≥ 1M<br>(≥ 1M, ≥ 1M) | ≥ 1M<br>(≥ 1M, ≥ 1M) | ≥ 1M<br>(≥ 1M, ≥ 1M) | ≥ 1M<br>(≥ 1M, ≥ 1M) | ≥ 1M<br>(≥ 1M, ≥ 1M) | ≥ 1M<br>(≥ 1M, ≥ 1M) |
| *Guangdong* | 0.0001%<br>(0.0000%, 0.0001%) | ≥ 1M<br>(≥ 1M, ≥ 1M) | ≥ 1M<br>(≥ 1M, ≥ 1M) | ≥ 1M<br>(≥ 1M, ≥ 1M) | ≥ 1M<br>(≥ 1M, ≥ 1M) | ≥ 1M<br>(≥ 1M, ≥ 1M) | ≥ 1M<br>(≥ 1M, ≥ 1M) |
| *Guangxi* | 0.0005%<br>(0.0003%, 0.0008%) | 676,130<br>(391,913, ≥ 1M) | ≥ 1M<br>(≥ 1M, ≥ 1M) | ≥ 1M<br>(≥ 1M, ≥ 1M) | ≥ 1M<br>(≥ 1M, ≥ 1M) | ≥ 1M<br>(≥ 1M, ≥ 1M) | ≥ 1M<br>(≥ 1M, ≥ 1M) |
| *Guizhou* | 0.0000%<br>(0.0000%, 0.0000%) | ≥ 1M<br>(≥ 1M, ≥ 1M) | ≥ 1M<br>(≥ 1M, ≥ 1M) | ≥ 1M<br>(≥ 1M, ≥ 1M) | ≥ 1M<br>(≥ 1M, ≥ 1M) | ≥ 1M<br>(≥ 1M, ≥ 1M) | ≥ 1M<br>(≥ 1M, ≥ 1M) |
| *Hainan* | 0.0000%<br>(0.0000%, 0.0000%) | Inf<br>(≥ 1M, Inf) | Inf<br>(≥ 1M, Inf) | Inf<br>(≥ 1M, Inf) | Inf<br>(≥ 1M, Inf) | Inf<br>(≥ 1M, Inf) | Inf<br>(≥ 1M, Inf) |
| *Hebei* | 0.0000%<br>(0.0000%, 0.0001%) | ≥ 1M<br>(≥ 1M, ≥ 1M) | ≥ 1M<br>(≥ 1M, ≥ 1M) | ≥ 1M<br>(≥ 1M, ≥ 1M) | ≥ 1M<br>(≥ 1M, ≥ 1M) | ≥ 1M<br>(≥ 1M, ≥ 1M) | ≥ 1M<br>(≥ 1M, ≥ 1M) |
| *Heilongjiang* | 0.0004%<br>(0.0002%, 0.0007%) | 726,439<br>(433,358, ≥ 1M) | ≥ 1M<br>(≥ 1M, ≥ 1M) | ≥ 1M<br>(≥ 1M, ≥ 1M) | ≥ 1M<br>(≥ 1M, ≥ 1M) | ≥ 1M<br>(≥ 1M, ≥ 1M) | ≥ 1M<br>(≥ 1M, ≥ 1M) |
| *Henan* | 0.0003%<br>(0.0002%, 0.0005%) | 969,289<br>(612,532, ≥ 1M) | ≥ 1M<br>(≥ 1M, ≥ 1M) | ≥ 1M<br>(≥ 1M, ≥ 1M) | ≥ 1M<br>(≥ 1M, ≥ 1M) | ≥ 1M<br>(≥ 1M, ≥ 1M) | ≥ 1M<br>(≥ 1M, ≥ 1M) |
| *Hubei* | 0.0000%<br>(0.0000%, 0.0000%) | ≥ 1M<br>(≥ 1M, ≥ 1M) | ≥ 1M<br>(≥ 1M, ≥ 1M) | ≥ 1M<br>(≥ 1M, ≥ 1M) | ≥ 1M<br>(≥ 1M, ≥ 1M) | ≥ 1M<br>(≥ 1M, ≥ 1M) | ≥ 1M<br>(≥ 1M, ≥ 1M) |
| *Hunan* | 0.0000%<br>(0.0000%, 0.0000%) | ≥ 1M<br>(≥ 1M, ≥ 1M) | ≥ 1M<br>(≥ 1M, ≥ 1M) | ≥ 1M<br>(≥ 1M, ≥ 1M) | ≥ 1M<br>(≥ 1M, ≥ 1M) | ≥ 1M<br>(≥ 1M, ≥ 1M) | ≥ 1M<br>(≥ 1M, ≥ 1M) |
| *Inner Mongolia* | 0.0000%<br>(0.0000%, 0.0002%) | ≥ 1M<br>(≥ 1M, ≥ 1M) | ≥ 1M<br>(≥ 1M, ≥ 1M) | ≥ 1M<br>(≥ 1M, ≥ 1M) | ≥ 1M<br>(≥ 1M, ≥ 1M) | ≥ 1M<br>(≥ 1M, ≥ 1M) | ≥ 1M<br>(≥ 1M, ≥ 1M) |
| *Jiangsu* | 0.0006%<br>(0.0004%, 0.0011%) | 550,222<br>(267,476, 848,138) | ≥ 1M<br>(745,755, ≥ 1M) | ≥ 1M<br>(≥ 1M, ≥ 1M) | ≥ 1M<br>(≥ 1M, ≥ 1M) | ≥ 1M<br>(≥ 1M, ≥ 1M) | ≥ 1M<br>(≥ 1M, ≥ 1M) |
| *Jiangxi* | 0.0000%<br>(0.0000%, 0.0000%) | ≥ 1M<br>(≥ 1M, ≥ 1M) | ≥ 1M<br>(≥ 1M, ≥ 1M) | ≥ 1M<br>(≥ 1M, ≥ 1M) | ≥ 1M<br>(≥ 1M, ≥ 1M) | ≥ 1M<br>(≥ 1M, ≥ 1M) | ≥ 1M<br>(≥ 1M, ≥ 1M) |
| *Jilin* | 0.0000%<br>(0.0000%, 0.0000%) | ≥ 1M<br>(≥ 1M, ≥ 1M) | ≥ 1M<br>(≥ 1M, ≥ 1M) | ≥ 1M<br>(≥ 1M, ≥ 1M) | ≥ 1M<br>(≥ 1M, ≥ 1M) | ≥ 1M<br>(≥ 1M, ≥ 1M) | ≥ 1M<br>(≥ 1M, ≥ 1M) |
| *Liaoning* | 0.0016%<br>(0.0008%, 0.0028%) | 189,757<br>(111,558, 395,974) | 529,065<br>(311,038, ≥ 1M) | ≥ 1M<br>(≥ 1M, ≥ 1M) | ≥ 1M<br>(≥ 1M, ≥ 1M) | ≥ 1M<br>(≥ 1M, ≥ 1M) | ≥ 1M<br>(826,341, ≥ 1M) |
| *Ningxia* | 0.0000%<br>(0.0000%, 0.0001%) | ≥ 1M<br>(≥ 1M, ≥ 1M) | ≥ 1M<br>(≥ 1M, ≥ 1M) | ≥ 1M<br>(≥ 1M, ≥ 1M) | ≥ 1M<br>(≥ 1M, ≥ 1M) | ≥ 1M<br>(≥ 1M, ≥ 1M) | ≥ 1M<br>(≥ 1M, ≥ 1M) |
| *Qinghai* | 0.0000%<br>(0.0000%, 0.0001%) | ≥ 1M<br>(≥ 1M, ≥ 1M) | ≥ 1M<br>(≥ 1M, ≥ 1M) | ≥ 1M<br>(≥ 1M, ≥ 1M) | ≥ 1M<br>(≥ 1M, ≥ 1M) | ≥ 1M<br>(≥ 1M, ≥ 1M) | ≥ 1M<br>(≥ 1M, ≥ 1M) |
| *Shaanxi* | 0.0000%<br>(0.0000%, 0.0001%) | ≥ 1M<br>(≥ 1M, ≥ 1M) | ≥ 1M<br>(≥ 1M, ≥ 1M) | ≥ 1M<br>(≥ 1M, ≥ 1M) | ≥ 1M<br>(≥ 1M, ≥ 1M) | ≥ 1M<br>(≥ 1M, ≥ 1M) | ≥ 1M<br>(≥ 1M, ≥ 1M) |
| *Shandong* | 0.0001%<br>(0.0001%, 0.0002%) | ≥ 1M<br>(≥ 1M, ≥ 1M) | ≥ 1M<br>(≥ 1M, ≥ 1M) | ≥ 1M<br>(≥ 1M, ≥ 1M) | ≥ 1M<br>(≥ 1M, ≥ 1M) | ≥ 1M<br>(≥ 1M, ≥ 1M) | ≥ 1M<br>(≥ 1M, ≥ 1M) |
| *Shanghai* | 0.0009%<br>(0.0005%, 0.0014%) | 353,414<br>(217,455, 664,360) | 985,361<br>(606,290, ≥ 1M) | ≥ 1M<br>(≥ 1M, ≥ 1M) | ≥ 1M<br>(≥ 1M, ≥ 1M) | ≥ 1M<br>(≥ 1M, ≥ 1M) | ≥ 1M<br>(≥ 1M, ≥ 1M) |
| *Shanxi* | 0.0000%<br>(0.0000%, 0.0000%) | ≥ 1M<br>(≥ 1M, ≥ 1M) | ≥ 1M<br>(≥ 1M, ≥ 1M) | ≥ 1M<br>(≥ 1M, ≥ 1M) | ≥ 1M<br>(≥ 1M, ≥ 1M) | ≥ 1M<br>(≥ 1M, ≥ 1M) | ≥ 1M<br>(≥ 1M, ≥ 1M) |
| *Sichuan* | 0.0001%<br>(0.0000%, 0.0002%) | ≥ 1M<br>(≥ 1M, ≥ 1M) | ≥ 1M<br>(≥ 1M, ≥ 1M) | ≥ 1M<br>(≥ 1M, ≥ 1M) | ≥ 1M<br>(≥ 1M, ≥ 1M) | ≥ 1M<br>(≥ 1M, ≥ 1M) | ≥ 1M<br>(≥ 1M, ≥ 1M) |
| *Tianjin* | 0.001%<br>(0.0003%, 0.003%) | 315,434<br>(101,042, 939,120) | 879,469<br>(281,716, ≥ 1M) | ≥ 1M<br>(≥ 1M, ≥ 1M) | ≥ 1M<br>(≥ 1M, ≥ 1M) | ≥ 1M<br>(≥ 1M, ≥ 1M) | ≥ 1M<br>(748,440, ≥ 1M) |
| *Tibet* | *n.d.* | --- | --- | --- | --- | --- | --- |
| *Xinjiang* | 0.0000%<br>(0.0000%, 0.0000%) | ≥ 1M<br>(≥ 1M, Inf) | ≥ 1M<br>(≥ 1M, Inf) | ≥ 1M<br>(≥ 1M, Inf) | ≥ 1M<br>(≥ 1M, Inf) | ≥ 1M<br>(≥ 1M, Inf) | ≥ 1M<br>(≥ 1M, Inf) |
| *Yunnan* | 0.0009%<br>(0.0005%, 0.0016%) | 350,800<br>(190,158, 584,150) | 978,072<br>(530,183, ≥ 1M) | ≥ 1M<br>(≥ 1M, ≥ 1M) | ≥ 1M<br>(≥ 1M, ≥ 1M) | ≥ 1M<br>(≥ 1M, ≥ 1M) | ≥ 1M<br>(≥ 1M, ≥ 1M) |
| *Zhejiang* | 0.0000%<br>(0.0000%, 0.0000%) | ≥ 1M<br>(≥ 1M, ≥ 1M) | ≥ 1M<br>(≥ 1M, ≥ 1M) | ≥ 1M<br>(≥ 1M, ≥ 1M) | ≥ 1M<br>(≥ 1M, ≥ 1M) | ≥ 1M<br>(≥ 1M, ≥ 1M) | ≥ 1M<br>(≥ 1M, ≥ 1M) |
| **Egypt** | 0.0726%<br>(0.0273%, 0.1138%) | 4,226<br>(2,697, 11,256) | 11,781<br>(7,518, 31,382) | 45,099<br>(28,780, 120,129) | 47,661<br>(30,415, 126,955) | 46,602<br>(29,740, 124,133) | 31,300<br>(19,974, 83,373) |
| **European Union** | 3.9648%<br>(3.0069%, 5.4727%) | 77<br>(56, 102) | 216<br>(156, 285) | 826<br>(598, 1,089) | 873<br>(632, 1,151) | 854<br>(618, 1,125) | 573<br>(415, 756) |
| *Austria* | 7.582%<br>(7.2889%, 9.6649%) | 40<br>(32, 42) | 113<br>(89, 117) | 432<br>(339, 449) | 456<br>(358, 475) | 446<br>(350, 464) | 300<br>(235, 312) |
| *Belgium* | 6.6396%<br>(4.5097%, 8.7201%) | 46<br>(35, 68) | 129<br>(98, 190) | 493<br>(376, 726) | 521<br>(397, 767) | 510<br>(388, 750) | 342<br>(261, 504) |
| *Bulgaria* | 2.2831%<br>(1.5615%, 3.1896%) | 134<br>(96, 197) | 375<br>(268, 548) | 1,434<br>(1,027, 2,097) | 1,516<br>(1,085, 2,217) | 1,482<br>(1,061, 2,167) | 996<br>(713, 1,456) |
| *Croatia* | 6.2509%<br>(5.4277%, 7.3272%) | 49<br>(42, 57) | 137<br>(117, 158) | 524<br>(447, 603) | 554<br>(472, 638) | 541<br>(462, 623) | 364<br>(310, 419) |
| *Cyprus* | 1.8986% | 162 | 451 | 1,725 | 1,823 | 1,782 | 1,197 |
|  | (1.3388%, 2.5765%) | (119, 229) | (332, 639) | (1,271, 2,446) | (1,343, 2,585) | (1,313, 2,528) | (882, 1,698) |
| *Czech Republic* | 10.128% | 30 | 84 | 323 | 342 | 334 | 224 |
|  | (9.0398%, 10.9588%) | (28, 34) | (78, 95) | (299, 362) | (316, 383) | (309, 374) | (207, 251) |
| *Denmark* | 4.4201% | 69 | 194 | 741 | 783 | 766 | 514 |
|  | (3.237%, 5.9783%) | (51, 95) | (143, 264) | (548, 1,012) | (579, 1,069) | (566, 1,045) | (380, 702) |
| *Estonia* | 3.2309% | 95 | 265 | 1,014 | 1,071 | 1,047 | 703 |
|  | (2.4958%, 4.555%) | (67, 123) | (188, 343) | (719, 1,312) | (760, 1,387) | (743, 1,356) | (499, 911) |
| *Finland* | 1.7646% | 174 | 485 | 1,856 | 1,961 | 1,918 | 1,288 |
|  | (1.0067%, 3.4921%) | (88, 305) | (245, 850) | (938, 3,253) | (991, 3,438) | (969, 3,361) | (651, 2,258) |
| *France* | 0.8692% | 353 | 984 | 3,768 | 3,982 | 3,893 | 2,615 |
|  | (0.3183%, 1.6579%) | (185, 964) | (516, 2,688) | (1,975, 10,289) | (2,088, 10,874) | (2,041, 10,632) | (1,371, 7,141) |
| *Germany* | 2.9448% | 104 | 291 | 1,112 | 1,175 | 1,149 | 772 |
|  | (2.2657%, 7.0048%) | (44, 135) | (122, 378) | (468, 1,445) | (494, 1,528) | (483, 1,494) | (324, 1,003) |
| *Greece* | 3.4772% | 88 | 246 | 942 | 995 | 973 | 654 |
|  | (2.5819%, 5.7358%) | (53, 119) | (149, 331) | (571, 1,268) | (603, 1,340) | (590, 1,311) | (396, 880) |
| *Hungary* | 5.4027% | 57 | 158 | 606 | 641 | 626 | 421 |
|  | (3.4865%, 8.0329%) | (38, 88) | (107, 245) | (408, 939) | (431, 993) | (421, 971) | (283, 652) |
| *Ireland* | 2.7065% | 113 | 316 | 1,210 | 1,279 | 1,250 | 840 |
|  | (1.3036%, 4.41%) | (70, 235) | (194, 656) | (743, 2,512) | (785, 2,655) | (767, 2,596) | (515, 1,744) |
| *Italy* | 0.8517% | 360 | 1,005 | 3,845 | 4,064 | 3,973 | 2,669 |
|  | (0.6251%, 1.2349%) | (248, 491) | (693, 1,369) | (2,652, 5,239) | (2,803, 5,537) | (2,740, 5,414) | (1,841, 3,636) |
| *Latvia* | 4.7632% | 64 | 180 | 688 | 727 | 710 | 477 |
|  | (3.2874%, 6.8327%) | (45, 93) | (125, 260) | (479, 996) | (507, 1,053) | (495, 1,029) | (333, 691) |
| *Lithuania* | 3.1464% | 98 | 272 | 1,041 | 1,100 | 1,076 | 722 |
|  | (2.4847%, 4.4227%) | (69, 123) | (193, 344) | (740, 1,318) | (783, 1,393) | (765, 1,362) | (514, 915) |
| *Luxembourg* | 1.7269% | 178 | 495 | 1,896 | 2,004 | 1,960 | 1,316 |
|  | (1.211%, 2.6969%) | (114, 253) | (317, 706) | (1,214, 2,704) | (1,283, 2,858) | (1,255, 2,794) | (843, 1,877) |
| *Malta* | 0.9345% | 328 | 915 | 3,504 | 3,703 | 3,621 | 2,432 |
|  | (0.6545%, 1.2744%) | (241, 469) | (671, 1,307) | (2,570, 5,004) | (2,716, 5,288) | (2,655, 5,171) | (1,783, 3,473) |
| *Netherlands* | 6.4673% | 47 | 132 | 506 | 535 | 523 | 351 |
|  | (4.752%, 8.3297%) | (37, 65) | (103, 180) | (393, 689) | (416, 728) | (406, 712) | (273, 478) |
| *Poland* | 2.118% | 145 | 404 | 1,546 | 1,634 | 1,598 | 1,073 |
|  | (1.9921%, 2.6347%) | (116, 154) | (325, 429) | (1,243, 1,644) | (1,314, 1,737) | (1,284, 1,699) | (863, 1,141) |
| *Portugal* | 0.5193% | 591 | 1,647 | 6,306 | 6,665 | 6,516 | 4,377 |
|  | (0.2801%, 0.7387%) | (415, 1,096) | (1,158, 3,055) | (4,433, 1,1693) | (4,685, 12,358) | (4,581, 12,083) | (3,077, 8,115) |
| *Romania* | 4.4927% | 68 | 190 | 729 | 770 | 753 | 506 |
|  | (2.8969%, 6.0855%) | (50, 106) | (141, 295) | (538, 1,131) | (569, 1,195) | (556, 1,168) | (373, 785) |
| *Slovakia* | 9.1138% | 34 | 94 | 359 | 380 | 371 | 249 |
|  | (7.8365%, 10.6227%) | (29, 39) | (81, 109) | (308, 418) | (326, 442) | (319, 432) | (214, 290) |
| *Slovenia* | 9.2961% | 33 | 92 | 352 | 372 | 364 | 244 |
|  | (6.2612%, 14.0264%) | (22, 49) | (61, 137) | (233, 523) | (247, 553) | (241, 540) | (162, 363) |
| *Spain* | 0.0575% | 5,333 | 14,870 | 56,923 | 60,157 | 58,821 | 39,506 |
|  | (0.0369%, 0.0874%) | (3,509, 8,315) | (9,785, 23,183) | (37,455, 88,744) | (39,584, 93,786) | (38,704, 91,702) | (25,995, 61,591) |
| *Sweden* | *n.d.* | --- | --- | --- | --- | --- | --- |
| **Iceland** | 2.197% | 140 | 389 | 1,491 | 1,575 | 1,540 | 1,035 |
|  | (1.3208%, 4.0252%) | (76, 232) | (213, 648) | (814, 2,480) | (860, 2,620) | (841, 2,562) | (565, 1,721) |
| **India** | 0.0904% | 3,395 | 9,466 | 36,235 | 38,294 | 37,443 | 25,148 |
|  | (0.0789%, 0.2077%) | (1,477, 3,887) | (4,119, 10,836) | (15,769, 41,482) | (16,665, 43,838) | (16,294, 42,864) | (10,944, 28,789) |
| **Indonesia** | 0.0101% | 30,352 | 84,624 | 323,936 | 342,341 | 334,734 | 224,821 |
|  | (0.005%, 0.0217%) | (14,139, 61,685) | (39,422, 171,986) | (150,906, 658,356) | (159,480, 695,762) | (155,936, 680,301) | (104,733, 456,918) |
| **Israel** | 0.1221% | 2,513 | 7,007 | 26,821 | 28,345 | 27,715 | 18,615 |
|  | (0.0792%, 0.1805%) | (1,700, 3,876) | (4,740, 10,806) | (18,144, 41,365) | (19,175, 43,716) | (18,749, 42,744) | (12,592, 28,709) |
| **Japan** | 0.0056% | 55,018 | 153,397 | 587,196 | 620,559 | 606,769 | 407,531 |
|  | (0.0034%, 0.0119%) | (25,686, 90,788) | (71,615, 253,127) | (274,139, 968,960) | (289,715, ≥ 1M) | (283,277, ≥ 1M) | (190,261, 672,487) |
| **Mexico** | 0.2971% | 1,033 | 2,879 | 11,022 | 11,648 | 11,389 | 7,649 |
|  | (0.1372%, 0.68%) | (451, 2,236) | (1,258, 6,235) | (4,816, 23,867) | (5,090, 25,223) | (4,977, 24,663) | (3,343, 16,565) |
| **New Zealand** | 0.1437% | 2,136 | 5,955 | 22,795 | 24,090 | 23,555 | 15,820 |
|  | (0.1056%, 0.2102%) | (1,460, 2,907) | (4,070, 8,105) | (15,578, 31,027) | (16,463, 32,790) | (16,097, 32,061) | (10,812, 21,534) |
| **Nigeria** | 0.0051% | 60,720 | 169,295 | 648,053 | 684,874 | 669,655 | 449,768 |
|  | (0.0025%, 0.0109%) | (28,140, 124,082) | (78,459, 345,955) | (300,336, ≥ 1M) | (317,400, ≥ 1M) | (310,347, ≥ 1M) | (208,442, 919,104) |
| **Norway** | 2.7376% | 112 | 313 | 1,196 | 1,264 | 1,236 | 830 |
|  | (2.0521%, 4.0047%) | (77, 150) | (214, 417) | (818, 1,596) | (864, 1,687) | (845, 1,649) | (568, 1,108) |
| **Qatar** | 0.4208% | 729 | 2,033 | 7,783 | 8,226 | 8,043 | 5,402 |
|  | (0.236%, 0.5729%) | (536, 1,300) | (1,493, 3,626) | (5,716, 13,879) | (6,041, 14,668) | (5,907, 14,342) | (3,967, 9,633) |
| **Russia** | 0.7135% | 430 | 1,199 | 4,590 | 4,850 | 4,743 | 3,185 |
|  | (0.6167%, 0.8016%) | (383, 498) | (1,067, 1,387) | (4,086, 5,310) | (4,318, 5,612) | (4,222, 5,487) | (2,836, 3,686) |
| **Saudi Arabia** | 0.002% | 150,840 | 420,561 | ≥ 1M | ≥ 1M | ≥ 1M | ≥ 1M |
|  | (0.0006%, 0.0052%) | (59,130, 482,837) | (164,862, ≥ 1M) | (631,084, ≥ 1M) | (666,941, ≥ 1M) | (652,121, ≥ 1M) | (437,991, ≥ 1M) |
| **Serbia** | 1.3359% | 230 | 640 | 2,451 | 2,591 | 2,533 | 1,701 |
|  | (0.9266%, 2.0453%) | (150, 331) | (418, 923) | (1,601, 3,534) | (1,692, 3,735) | (1,655, 3,652) | (1,111, 2,453) |
| **South Africa** | 0.048% | 6,399 | 17,841 | 68,293 | 72,173 | 70,569 | 47,397 |
|  | (0.025%, 0.1222%) | (2,511, 12,254) | (7,002, 34,164) | (26,803, 130,780) | (28,326, 138,210) | (27,697, 135,139) | (18,602, 90,765) |
| **South Korea** | 0.3687% | 832 | 2,320 | 8,881 | 9,386 | 9,177 | 6,164 |
|  | (0.2869%, 0.6332%) | (485, 1,069) | (1,351, 2,982) | (5,172, 11,414) | (5,466, 12,063) | (5,345, 11,795) | (3,590, 7,922) |
| **Switzerland** | 1.2712% | 241 | 673 | 2,576 | 2,723 | 2,662 | 1,788 |
|  | (0.9103%, 2.7461%) | (112, 337) | (312, 940) | (1,193, 3,597) | (1,260, 3,802) | (1,232, 3,717) | (828, 2,497) |
| **Turkey** | 0.5889% | 521 | 1,453 | 5,561 | 5,877 | 5,747 | 3,860 |
|  | (0.495%, 0.6974%) | (440, 620) | (1,227, 1,728) | (4,696, 6,616) | (4,962, 6,992) | (4,852, 6,836) | (3,259, 4,592) |
| **Ukraine** | 4.3754% | 70 | 196 | 748 | 791 | 773 | 519 |
|  | (2.9122%, 5.3887%) | (57, 105) | (159, 294) | (608, 1,125) | (642, 1,188) | (628, 1,162) | (422, 780) |
| **United Arab Emirates** | 0.094% | 3,265 | 9,103 | 34,847 | 36,827 | 36,009 | 24,185 |
|  | (0.0494%, 0.2498%) | (1,229, 6,211) | (3,426, 17,316) | (13,113, 66,286) | (13,858, 70,053) | (13,550, 68,496) | (9,101, 46,005) |
| **United Kingdom** | 2.5715% | 119 | 333 | 1,274 | 1,346 | 1,316 | 884 |
|  | (2.0297%, 3.4445%) | (89, 151) | (248, 421) | (951, 1,613) | (1,005, 1,705) | (982, 1,667) | (660, 1,120) |
| **United States of America** | 1.7919% | 171 | 477 | 1,828 | 1,931 | 1,889 | 1,268 |
|  | (1.1619%, 2.687%) | (114, 264) | (318, 736) | (1,219, 2,819) | (1,288, 2,979) | (1,259, 2,912) | (846, 1,956) |
| *Alabama* | 1.6329% | 188 | 524 | 2,006 | 2,120 | 2,072 | 1,392 |
|  | (0.9706%, 2.5944%) | (118, 316) | (330, 881) | (1,262, 3,374) | (1,334, 3,566) | (1,304, 3,487) | (876, 2,342) |
| *Alaska* | 3.9535% | 78 | 216 | 828 | 875 | 856 | 575 |
|  | (2.5054%, 5.87%) | (52, 122) | (146, 341) | (558, 1,307) | (590, 1,381) | (577, 1,351) | (387, 907) |
| Arizona | 2.7643% | 111 | 309 | 1,185 | 1,252 | 1,224 | 822 |
|  | (1.8833%, 3.7897%) | (81, 163) | (226, 454) | (864, 1,739) | (913, 1,838) | (893, 1,797) | (600, 1,207) |
| Arkansas | 0.5864% | 523 | 1,459 | 5,585 | 5,902 | 5,771 | 3,876 |
|  | (0.344%, 0.9375%) | (327, 892) | (913, 2,487) | (3,493, 9,519) | (3,692, 10,060) | (3,610, 9,836) | (2,424, 6,606) |
| California | 1.9291% | 159 | 443 | 1,698 | 1,794 | 1,754 | 1,178 |
|  | (1.1926%, 2.9288%) | (105, 257) | (292, 717) | (1,118, 2,746) | (1,182, 2,902) | (1,155, 2838) | (776, 1,906) |
| Colorado | 3.8648% | 79 | 221 | 847 | 896 | 876 | 588 |
|  | (2.5974%, 5.5929%) | (55, 118) | (153, 329) | (586, 1,261) | (619, 1,333) | (605, 1,303) | (406, 875) |
| Connecticut | 0.7765% | 395 | 1,102 | 4,218 | 4,457 | 4,358 | 2,927 |
|  | (0.3348%, 1.3932%) | (220, 917) | (614, 2,555) | (2,351, 9,782) | (2,484, 10,338) | (2,429, 10,108) | (1,631, 6,789) |
| Delaware | 1.4611% | 210 | 586 | 2,241 | 2,369 | 2,316 | 1,556 |
|  | (0.8877%, 2.1985%) | (140, 346) | (389, 964) | (1,490, 3,689) | (1,574, 3,899) | (1,539, 3,812) | (1,034, 2,560) |
| District of Columbia | 0.9928% | 309 | 862 | 3,299 | 3,486 | 3,409 | 2,289 |
|  | (0.4965%, 1.8429%) | (166, 618) | (464, 1,723) | (1,777, 6,596) | (1,878, 6,971) | (1,836, 6,816) | (1,233, 4,578) |
| Florida | 0.1491% | 2,058 | 5,739 | 21,969 | 23,218 | 22,702 | 15,247 |
|  | (0.0735%, 0.3026%) | (1,014, 4,177) | (2,828, 11,646) | (10,824, 44,579) | (11,439, 47,112) | (11,185, 46,065) | (7,512, 30,939) |
| Georgia | 0.3715% | 826 | 2,303 | 8,816 | 9,317 | 9,110 | 6,119 |
|  | (0.1839%, 0.7033%) | (436, 1,668) | (1,216, 4,651) | (4,656, 17,804) | (4,921, 18,815) | (4,811, 18,397) | (3,232, 12,356) |
| Hawaii | 1.1000% | 279 | 778 | 2,977 | 3,146 | 3,076 | 2,066 |
|  | (0.6391%, 1.6064%) | (191, 480) | (533, 1,339) | (2,039, 5,125) | (2,155, 5,416) | (2,107, 5,295) | (1,415, 3,557) |
| Idaho | 1.7874% | 172 | 479 | 1,832 | 1,936 | 1,893 | 1,272 |
|  | (1.1244%, 2.779%) | (110, 273) | (308, 761) | (1,178, 2,913) | (1,245, 3,078) | (1,218, 3,010) | (818, 2,021) |
| Illinois | 1.6601% | 185 | 515 | 1,973 | 2,085 | 2,039 | 1,369 |
|  | (1.034%, 2.4965%) | (123, 297) | (343, 827) | (1,312, 3,167) | (1,386, 3,347) | (1,356, 3,273) | (910, 2,198) |
| Indiana | 1.887% | 163 | 453 | 1,736 | 1,834 | 1,793 | 1,204 |
|  | (1.2236%, 2.7637%) | (111, 251) | (310, 699) | (1,185, 2,677) | (1,252, 2,829) | (1,224, 2,766) | (822, 1,858) |
| Iowa | 3.1211% | 98 | 274 | 1,049 | 1,109 | 1,084 | 728 |
|  | (2.073%, 4.6638%) | (66, 148) | (183, 413) | (702, 1,580) | (742, 1,670) | (726, 1,633) | (487, 1,096) |
| Kansas | 2.205% | 139 | 388 | 1,485 | 1,570 | 1,535 | 1,031 |
|  | (1.4071%, 3.3584%) | (91, 218) | (255, 608) | (975, 2,327) | (1,031, 2,460) | (1,008, 2,405) | (677, 1,615) |
| Kentucky | 1.3104% | 234 | 653 | 2,499 | 2,641 | 2,583 | 1,735 |
|  | (0.7906%, 2.1014%) | (146, 388) | (407, 1,082) | (1,558, 4,142) | (1,647, 4,377) | (1,610, 4,280) | (1,082, 2,875) |
| Louisiana | 0.1816% | 1,689 | 4,710 | 18,029 | 19,053 | 18,630 | 12,513 |
|  | (0.0994%, 0.3329%) | (922, 3,086) | (2,570, 8,604) | (9,838, 32,934) | (10,397, 34,805) | (10,166, 34,032) | (6,828, 22,857) |
| Maine | 4.8815% | 63 | 175 | 671 | 709 | 693 | 466 |
|  | (3.4574%, 6.8067%) | (45, 89) | (126, 247) | (481, 947) | (508, 1,001) | (497, 979) | (334, 657) |
| Maryland | 1.2408% | 247 | 689 | 2,639 | 2,789 | 2,727 | 1,832 |
|  | (0.7756%, 1.9289%) | (159, 396) | (444, 1,103) | (1,698, 4,223) | (1,794, 4,463) | (1,754, 4,363) | (1,178, 2,931) |
| Massachusetts | 1.5626% | 196 | 547 | 2,096 | 2,215 | 2,166 | 1,455 |
|  | (0.995%, 2.3719%) | (129, 308) | (361, 860) | (1,381, 3,291) | (1,459, 3,478) | (1,427, 3,401) | (958, 2,284) |
| Michigan | 3.122% | 98 | 274 | 1,049 | 1,109 | 1,084 | 728 |
|  | (2.3887%, 4.1254%) | (74, 128) | (207, 358) | (794, 1,371) | (839, 1,449) | (820, 1,417) | (551, 952) |
| Minnesota | 3.7262% | 82 | 230 | 879 | 929 | 908 | 610 |
|  | (2.5439%, 5.2348%) | (59, 121) | (163, 336) | (626, 1,287) | (661, 1,360) | (646, 1,330) | (434, 893) |
| *Mississippi* | 0.3868% | 793 | 2,212 | 8,467 | 8,948 | 8,749 | 5,876 |
|  | (0.2061%, 0.6979%) | (440, 1,489) | (1,226, 4,152) | (4,692, 15,894) | (4,959, 16,797) | (4,849, 16,423) | (3,257, 11,031) |
| *Missouri* | 1.9784% | 155 | 432 | 1,655 | 1,749 | 1,711 | 1,149 |
|  | (1.3369%, 2.8753%) | (107, 230) | (298, 640) | (1,139, 2,450) | (1,204, 2,589) | (1,177, 2,531) | (790, 1,700) |
| *Montana* | 2.493% | 123 | 343 | 1,314 | 1,388 | 1,357 | 912 |
|  | (1.5002%, 3.9065%) | (79, 205) | (219, 570) | (838, 2,183) | (886, 2,307) | (866, 2,256) | (582, 1,515) |
| *Nebraska* | 2.8688% | 107 | 298 | 1,142 | 1,206 | 1,180 | 792 |
|  | (1.7603%, 4.6931%) | (65, 174) | (182, 486) | (698, 1,860) | (737, 1,966) | (721, 1,922) | (484, 1,291) |
| *Nevada* | 1.6236% | 189 | 527 | 2,017 | 2,132 | 2,084 | 1,400 |
|  | (0.9193%, 2.587%) | (119, 334) | (331, 931) | (1,266, 3,562) | (1,338, 3,765) | (1,308, 3,681) | (879, 2,472) |
| *New Hampshire* | 5.5646% | 55 | 154 | 589 | 622 | 608 | 408 |
|  | (3.4291%, 7.8332%) | (39, 89) | (109, 249) | (418, 955) | (442, 1,009) | (432, 987) | (290, 663) |
| *New Jersey* | 1.3089% | 234 | 654 | 2,502 | 2,644 | 2,585 | 1,736 |
|  | (0.9035%, 1.805%) | (170, 340) | (474, 947) | (1,814, 3,625) | (1,917, 3,831) | (1,875, 3,746) | (1,259, 2,516) |
| *New Mexico* | 4.3246% | 71 | 198 | 757 | 800 | 783 | 526 |
|  | (2.8308%, 6.1917%) | (50, 108) | (138, 302) | (529, 1,157) | (559, 1,223) | (547, 1,195) | (367, 803) |
| *New York* | 3.2186% | 95 | 266 | 1,018 | 1,075 | 1,051 | 706 |
|  | (2.3387%, 4.3285%) | (71, 131) | (198, 366) | (757, 1,400) | (800, 1,480) | (782, 1,447) | (525, 972) |
| *North Carolina* | 0.8165% | 376 | 1,048 | 4,011 | 4,239 | 4,145 | 2,784 |
|  | (0.4574%, 1.4275%) | (215, 671) | (599, 1,870) | (2,294, 7,160) | (2,424, 7,567) | (2,371, 7,399) | (1,592, 4,969) |
| *North Dakota* | 2.5186% | 122 | 340 | 1,300 | 1,374 | 1,344 | 902 |
|  | (1.6199%, 3.8533%) | (80, 189) | (222, 528) | (850, 2,022) | (898, 2,137) | (878, 2,089) | (590, 1,403) |
| *Ohio* | 2.4753% | 124 | 346 | 1,323 | 1,398 | 1,367 | 918 |
|  | (1.7355%, 3.4126%) | (90, 177) | (251, 493) | (960, 1,887) | (1,014, 1,994) | (992, 1,950) | (666, 1,310) |
| *Oklahoma* | 1.11% | 276 | 771 | 2,951 | 3,118 | 3,049 | 2,048 |
|  | (0.6703%, 1.785%) | (172, 458) | (479, 1,276) | (1,835, 4,886) | (1,939, 5,163) | (1,896, 5,049) | (1,273, 3,391) |
| *Oregon* | 1.938% | 158 | 441 | 1,690 | 1,786 | 1,746 | 1,173 |
|  | (1.114%, 3.1602%) | (97, 275) | (271, 768) | (1,036, 2,940) | (1,095, 3,107) | (1,071, 3,038) | (719, 2,040) |
| *Pennsylvania* | 3.1145% | 99 | 275 | 1,052 | 1,111 | 1,087 | 730 |
|  | (2.2874%, 4.1416%) | (74, 134) | (207, 374) | (791, 1,432) | (836, 1,513) | (817, 1,479) | (549, 994) |
| *Rhode Island* | 1.3159% | 233 | 650 | 2,489 | 2,630 | 2,572 | 1,727 |
|  | (0.8309%, 1.9954%) | (154, 369) | (429, 1,030) | (1,641, 3,941) | (1,735, 4,165) | (1,696, 4,073) | (1,139, 2,735) |
| *South Carolina* | 0.3772% | 814 | 2,268 | 8,683 | 9,177 | 8,973 | 6,026 |
|  | (0.1976%, 0.7019%) | (437, 1,553) | (1,219, 4,329) | (4,666, 16,571) | (4,931, 17,513) | (4,821, 17,124) | (3,238, 11,501) |
| *South Dakota* | 1.9808% | 155 | 432 | 1,653 | 1,747 | 1,708 | 1,147 |
|  | (1.2039%, 3.1375%) | (98, 255) | (273, 711) | (1,044, 2,720) | (1,103, 2,875) | (1,079, 2,811) | (724, 1,888) |
| *Tennessee* | 0.5464% | 562 | 1,566 | 5,993 | 6,334 | 6,193 | 4,159 |
|  | (0.2905%, 0.9501%) | (323, 1,056) | (900, 2,945) | (3,447, 11,274) | (3,643, 11,915) | (3,562, 11,650) | (2,392, 7,824) |
| *Texas* | 0.4587% | 669 | 1,865 | 7,140 | 7,546 | 7,378 | 4,956 |
|  | (0.2479%, 0.8396%) | (365, 1,238) | (1,019, 3,451) | (3,900, 13,212) | (4,122, 13,962) | (4,031, 13,652) | (2,707, 9,169) |
| *Utah* | 1.3594% | 226 | 629 | 2,409 | 2,546 | 2,489 | 1,672 |
|  | (0.906%, 1.9593%) | (157, 339) | (437, 944) | (1,671, 3,615) | (1,766, 3,820) | (1,727, 3,735) | (1,160, 2,509) |
| *Vermont* | 5.7049% | 54 | 150 | 574 | 607 | 593 | 398 |
|  | (4.053%, 7.6921%) | (40, 76) | (111, 211) | (426, 808) | (450, 854) | (440, 835) | (295, 561) |
| *Virginia* | 1.5061% | 204 | 568 | 2,174 | 2,298 | 2,247 | 1,509 |
|  | (0.8939%, 2.369%) | (130, 343) | (361, 957) | (1,382, 3,664) | (1,461, 3,872) | (1,429, 3,786) | (959, 2,543) |
| *Washington* | 1.5935% | 193 | 537 | 2,055 | 2,172 | 2,124 | 1,426 |
|  | (0.8836%, 2.6517%) | (116, 347) | (323, 968) | (1,235, 3,706) | (1,305, 3,917) | (1,276, 3,830) | (857, 2,572) |
| *West Virginia* | 2.2764% | 135 | 376 | 1,439 | 1,520 | 1,487 | 998 |
|  | (1.5275%, 3.2917%) | (93, 201) | (260, 560) | (995, 2,144) | (1,051, 2,266) | (1,028, 2,216) | (690, 1,488) |
| *Wisconsin* | 3.3275% | 92 | 257 | 984 | 1,040 | 1,017 | 683 |
|  | (2.256%, 4.7204%) | (65, 136) | (181, 379) | (694, 1,452) | (733, 1,534) | (717, 1,500) | (482, 1,007) |
| *Wyoming* | 2.2208% | 138 | 385 | 1,475 | 1,558 | 1,524 | 1,023 |
|  | (1.4641%, 3.2833%) | (93, 210) | (261, 584) | (997, 2,237) | (1,054, 2,364) | (1,031, 2,311) | (692, 1,552) |

95% CI=95% confidence interval. Brackets are the 95% confidence intervals (lower limit, upper limit). *n.d.*=no data. NNE=number needed to exclude. NNEs are color-coded by the lower limit of the 95% CI: yellow ( $\geq 1,000$ and $< 10,000$ ), orange ( $\geq 10,000$ and $< 100,000$ ), red ( $\geq 100,000$ and $< 1,000,000$ ), and black ( $\geq 1,000,000$ ). NNEs $\geq 1,000,000$ were truncated to $\geq 1M$ . †Estimates of the baseline infection risk (i.e., point-prevalence of infectious cases in the jurisdiction) were taken from a database (https://decision-support-tools.com/) developed and regularly updated by Defence Research and Development Canada (DRDC), an agency of the Canadian Department of National Defence. Baseline infection risks are shown at most to the fourth decimal place due to space limitations. A true 0% baseline infection risk can be identified in the table when the NNE is infinite (Inf) because that means there were no infectious cases (NNE=1/0=infinite).

## DISCUSSION

There were two main findings of our study. First, the NNEs suggest that at least 1,000 unvaccinated people likely need to be excluded to prevent one SARS-CoV-2 transmission event in most types of settings for many jurisdictions, notably Australia, California, Canada, China, France, Israel, and others. Second, the NNEs of almost every jurisdiction examined are well within the range of the NNTs of ASA in primary prevention of CVD (≥ 250 to 333). There are four main implications of our study.

First, the two conservative assumptions of the model mean that the calculated NNEs likely represent the *minimum* number of unvaccinated people needed to exclude to prevent one SARS-CoV-2 transmission event in each jurisdiction and type of setting. The true NNEs are likely much higher, especially in jurisdictions with higher levels of vaccine/natural immunity.

Second, while SARS-CoV-2 vaccines are beneficial, the high NNEs suggest that excluding unvaccinated people has negligible benefits for reducing SARS-CoV-2 transmissions in many jurisdictions across the globe. This is because unvaccinated people are likely *not* at significant risk – in absolute terms – of transmitting SARS-CoV-2 to others in most types of settings (as of mid-to-end November 2021). This is why so many unvaccinated people likely need to be excluded to prevent one transmission event. While a meta-analysis found that the RRR of vaccines on SARs is ∼41%,^3^ current baseline transmission risks in most settings are negligible, such that a 41% decrease of a negligible risk is still a negligible absolute risk difference between vaccinated vs. unvaccinated people. This does *not* mean vaccines are not beneficial for reducing transmission; on the contrary, a 41% RRR in SARs in a population with high vaccination levels will lower the burden of SARS-CoV-2. Rather, it means that excluding unvaccinated people on the basis of the *relative* risk difference between vaccinated vs. unvaccinated people is likely not justifiable because baseline transmission risks are negligible for most types of settings and jurisdictions.

Third, societies and policymakers should urgently consider the harms of exclusion via VMVP. This is because these NNEs suggest that exclusion may not be a proportionate response to the risk unvaccinated people pose to others. Just as ASA is not recommended for primary prevention of CVD because the harms outweigh the benefits (NNTs ≥ 250 to 333), the harms of excluding unvaccinated people via VMVP may outweigh the benefits (NNEs ≥ 250 to 333). The analogy to ASA is made not because the outcomes are the same. Clearly, they are not: one is a within-individual risk (e.g., myocardial infarction), the other is a between-individual or ‘externalized’ risk where one or more other people may be impacted (i.e., transmission event). Rather, we are drawing a comparison between a public health intervention (i.e., exclusion) and an intervention widely recognized in medicine as having limited benefit due to the negligible ARRs. Thus, while the outcomes are different, the NNTs of ASA in primary prevention of CVD provide a benchmark for interpreting the magnitude of the NNEs. Based on this benchmark, our study suggests that this public health intervention (exclusion) has limited benefit. Given that many jurisdictions have or are planning to implement exclusionary policies via VMVP, there is an urgent need to consider the harms in light of the limited benefits. At least four harms should be considered. First, there is the possibility of staffing shortages from the loss of unvaccinated healthcare workers, which may have downstream effects on patient care. Second, there is the harm of unemployment and – in some cases – unemployability if vaccines are a condition of employment or higher-education in most organizations. Third, this would mean unvaccinated people and their families will face financial hardship. This may have downstream effects on their mental/physical health. A fourth harm of exclusion is the creation of a class of citizens who are not allowed to fully participate in many areas of society.

Fourth, we suggest that the NNE and the baseline infection risk (i.e., the point-prevalence of infectious cases) be included as key indicators for determining the harms vs. benefits of VMVP. It compliments other indicators, such as hospital/ICU occupancy, the basic and effective reproduction number, SARS-CoV-2-related hospitalizations/deaths, and regional vaccination rates. Moreover, because the model can be updated to account for any future potentially dominant variants (e.g., Omicron), the NNE enables efficient monitoring of the benefits of exclusion in real-time as the pandemic changes and variants emerge. This will improve public health because the NNE provides societies and policymakers with an intuitive metric to monitor whether to implement exclusionary policies via VMVP and when to stop these policies.

These findings depend on the accuracy of the assumptions of the NNE model and the accuracy of baseline infection risks estimates. The first assumption was that the Delta variant correction factor (1.97) we applied to the robust estimates of the wild-type SARs was accurate. We discovered that this was an accurate but conservative assumption since the corrected robust SAR in households was 26% higher than what has been observed in real-world transmission studies of the Delta variant in unvaccinated index cases.^24-26^ The higher corrected SARs 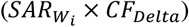 of the model increased the estimated baseline transmission risks, which lowered the NNEs. The second assumption was that the SARs for each type of setting are relatively consistent. This was supported by using robust estimates of the SARs from multiple meta-analyses. The third assumption was that the baseline infection risk is relatively stable across individuals in a population. This was addressed by calculating 95% CIs around the NNEs for each jurisdiction using the 95% CIs around the baseline infection risks. The fourth assumption was that (i) unvaccinated people have no natural immunity and (ii) the contacts of infected unvaccinated index cases have no vaccine or natural immunity. This was a strong conservative assumption because it means the estimated baseline transmission risks were higher than what would be predicted in a population with some level of vaccine/natural immunity because studies show that vaccine/natural immunity reduces SARs.^22 23^

Estimating the baseline infection risk in a population is challenging because it requires modeling various viral factors (e.g., incubation and infectious periods, asymptomatic cases) in addition to case counts to arrive at the current point-prevalence of infectious cases. There is likely some degree of underestimation of the baseline risk in some jurisdictions due to underreporting of cases. However, it is extremely difficult to accurately model the underreporting rate for each jurisdiction and how it changes over time because one is trying to model something where there is no data. This is shown by the extreme variation in underreporting estimates.^21 27^ Local context/knowledge remains essential for estimating underreporting rates in a jurisdiction over time. Since this is not available at a global scale, the degree of underestimation is unknown. Underestimation was partially addressed by utilizing the 95% CIs of the baseline infection risks in order to represent the likely *general range* of the baseline infection risks. The conservative assumptions also partially compensate for this because, as we just noted, they increased the estimated baseline transmission risks, which is what correcting for underreporting would do. Notwithstanding this limitation, we have provided a ‘proof of concept’ of how baseline infection risk data can be used to calculate the NNE to evaluate the benefits of excluding unvaccinated people to reduce SARS-CoV-2 transmissions.

Future research should focus on developing an online interactive global database of the NNEs and baseline infection risks in each region which is regularly updated. Furthermore, this interactive database should be linked to regional vaccination rates, SARS-CoV-2-related hospitalizations/deaths, hospital/ICU occupancy, and the basic and effective reproduction number to contextualize the NNEs within other important metrics. This would be helpful for societies and policymakers to evaluate in real-time the harms vs. benefits of excluding unvaccinated people. Health economic analyses are also needed to analyze the cost-effectiveness and cost-benefits of excluding unvaccinated people. Future studies could incorporate vaccine/natural immunity data into the model to explore the impact of herd immunity on the NNE. Finally, when new variants emerge (e.g., Omicron) and potentially outcompete the Delta variant, the NNEs should be updated with the transmissibility data of future dominant variants.

## Data Availability

The data used in this work are available at https://tinyurl.com/4m8mm4jh and https://decision-support-tools.com/.

## Acknowledgement

We thank numerous colleagues who provided helpful comments and criticisms during the writing of this study.

